# Estimating HPV16 genome copy number per infected cell in cervical smears

**DOI:** 10.1101/2024.06.13.24308781

**Authors:** Baptiste Elie, Vanina Boué, Philippe Paget-Bailly, Marie-Paule Algros, Alice Baraquin, Jean-Luc Prétet, Samuel Alizon, Nicolas Tessandier, Ignacio G. Bravo

## Abstract

Human papillomavirus (HPV) 16 is the most oncogenic biological agents for humans. However, essential quantitative aspects of its infection cycle remain inadequately characterized. Specifically, the proportion of infected cells and the viral copy number per infected cell in cervical smears are not well understood. To address this, we employed a combination of limiting dilution techniques and Bayesian statistics on routine cervical smears to estimate the frequency of infected cells and the viral copy number per cell. Our methodology was initially validated through numerical simulations and cell culture experiments. Subsequently, we analyzed 38 HPV16-positive cervical smears, comprising 26 samples from patients without cytological lesions and 12 from patients with low-grade lesions. Our findings indicated that the substantial variability in viral load across samples predominantly stemmed from differences in the frequency of infected cells. Additionally, the mean number of HPV copies per infected cell was consistently low across all samples, ranging from approximately 2.3 to 100 copies. However, in samples with low-grade lesionMarie-Paule Algross, this number was observed to double on average. These results challenge existing assumptions regarding the biology of HPV genital infections, which are typically asymptomatic or minimally symptomatic.

## Introduction

Human papillomaviruses (HPVs) are widespread and primarily cause asymptomatic and benign mucosal and skin infections [1]. However, specific oncogenic HPV genotypes are responsible for nearly all cervical cancers and for a significant proportion of anogenital and oropharyngeal cancers [2]. Cervical cancer prevention relies on two public health strategies: primary prevention through vaccination, targeting HPV genotypes responsible for at least 70% of cervical cancers, and secondary prevention through screening. Traditional screening focused on detecting precancerous and cancerous lesions, but there has been a shift towards molecular methods for detecting oncogenic HPVs, targetting viral DNA, RNA or assessing methylation status. Indeed, given the high prevalence of anogenital HPV infections, the mere detection of viral material in clinical samples often lacks specificity for cervical cancer risk [3]. Emerging screening methods that detect differential methylation levels in specific genomic regions show promise but may be difficult to implement for widespread use in resource-limited settings [4, 5]. Consequently, HPV viral genome load measurement presents a feasible and integrable option for enhancing cervical cancer triage algorithms [6].

Viral genome load levels vary among HPV genotypes for a given stage of infection [7]. Longitudinal studies have demonstrated a correlation between increased viral load and the risk of developing high-grade lesions, particularly for HPV16 and HPV18 [8, 9, 10, 11]. The detailed biological mechanisms linking variation in viral genome load and the natural history of the infection remain little studied. For instance, low-grade squamous intraepithelial lesions (LSIL) are associated with higher HPV16 viral loads compared to asymptomatic infections [12, 10]. However, there is conflicting evidence on whether LSIL can progress to high-grade lesions, with some studies showing an association [6] while others do not [13]. Understanding the biological mechanisms that explain the increased viral load associated with increasing cervical lesion severity is thus crucial.

In the human cervix, HPVs infect the basal layer of the pluristratified eptithelium, as well as the cervical transition zone between the monostratified endocervix and the pluristratified ectocervix. After cellular entry, the viral genome localises in the nucleus, where viral replication and transcription take place. The viral transcriptional repertoire changes in parallel with cell differentiation, from early open reading frames (ORFs) in the basal and parabasal layers to late ORFs (*L1* and *L2*) in differentiated keratinocytes, leading to capsid assembly, genome encapsidation, and virion production [1]. Additionally, despite recent advances in the development of murine models [14], the biology of asymptomatic genital infections, in particular from a quantitative perspective [15], remains poorly characterized [16]. Current understanding suggests that infected parabasal layers contain 100-1,000 viral genome copies per cell, increasing in the upper epithelial layers. Highly productive lesions in the rabbit papillomavirus model can reach millions of viral genome copies per cell in the upper layers [17], while high-grade human cervical lesions typically contain tens to hundreds of copies per cell [18, 19].

HPV genome load is typically expressed as the average number of viral genome copies per cell genome in the sample, which does not provide information about the frequency of infected cells or the “burst size” (*i*.*e*. the number of virions released per infected cell). HPV infections are non-cytolytic, and virions are released during keratinocyte desquamation. Estimating the “burst size” is challenging due to the loss of the nucleus in differentiating keratinocytes, which complicates further traditional quantification methods. Higher viral loads in lesion-derived samples compared to normal cytology samples may result from an increased frequency of infected cells, a higher number of viral copies per cell, or both. We implement an original approach that exploits broadly available asymptomatic cervical smears from screening leftovers. By combining the limiting dilution technique approach, widely used in HIV research [20] and other chronic viruses [21], with a Bayesian statistical framework [22], we simultaneously estimate the frequency of infected cells and the number of viral copies per cell. We also test the hypothesis that the higher viral genome load retrieved from LSIL compared to a normal cytology is related to an increase in the proportion of infected keratinocytes, in the mean number of viral copies per infected cell, or both.

## Methods

### Clinical samples

#### HPV16-positive cervical samples

Cervical samples were collected from women aged more than 18 attending the Department of Gynaecology and Obstetrics, Besanç on University Hospital (France). Cervical smears were obtained by scraping the cervix with a Rovers^™^ Cervex Brush^™^ and cells were secondarily discharged into the ThinPrep PreservCyt medium (Hologic). Samples were transferred to the laboratory and stored at room temperature until analysis using the Alinity m HR HPV Assay (Abbott) according to the manufacturer’s instructions. This assay allows the detection of 14 HPV genotypes, individually identifying HPV16, 18, and 45, and collectively identifying the other vaccine-preventable hrHPV genotypes (HPV 31, 33, 52, and 58) as group A, and the non-vaccine-preventable hrHPV (HPV35, 39, 51, 56, 59, 68 plus HPV66) as group B. Cytological results, expressed according to the Bethesda System [23], were obtained from medical records. All samples used were exclusively positive for the presence of HPV16 DNA.

#### HPV-negative cervical samples

Cervical smears without HPV16 infection used for protocol validation came from the study “Health in climacteric women belonging to NGOs. PAPILONG study”. Briefly, cervical samples were discharged into the ThinPrep PreservCyt medium (Hologic), cells pelleted by centrifugation and the DNA extracted using the DNAeasy kit (Qiagen). HPV genotyping was performed in duplicate using the INNO-LiPA HPV assay (Fujirebio, Les Ulis, France) following the manufacturer’s recommendations. This test, based on reverse hybridization after a polymerase chain reaction (PCR) step, allows type-specific detection of 32 viral genotypes within the Alphapapillomavirus genus, including oncogenic HPVs (HPV16, 18, 31, 33, 35, 39, 45, 51, 52, 56, 58, 59), possibly oncogenic (HPV68), probably oncogenic HPVs (HPV26, 53, 66, 67, 70, 73, 82), and non-oncogenic/unclassified HPVs (HPV6, 11, 40, 42, 43, 44, 54, 61, 62, 81, 83, 89). All samples used were negative for the presence of DNA of all genotypes tested.

### Cell cultures

The human SiHa cell lines, containing 1 to 3 HPV16 genome copies per cell, were obtained from ATCC (Manassas, VA, USA) and routinely tested for the presence of HPV. Cells were maintained at 37°C (5% CO2) in complete RPMI (Dutscher, Bernolsheim, France) or DMEM (Lonza, Levallois-Perret, France) supplemented with 10% fetal bovine serum (Thermo Fisher Scientific, Illkirch-Graffenstaden, France) and 10^5^U/L penicillin, 100mg/L streptomycin (Dutscher) in final concentration.

### Cell preparation and lysis

Cells conserved in Preservcyt (Hologic) were filtered with 70µm nylon cell strainer, pelleted via centrifugation at 600xg for 10 minutes, washed twice with Phosphate Buffer Saline (PBS, Corning), and resuspended in 100µL PBS. Cells were counted manually, with a phase-contrast microscope. Independent triplicate counts were performed and keratinocytes were distinguished from leukocytes based on shape and size.

Based on earlier results from high-grade lesions with low productivity [18, 19], we expected a low “burst size”. Therefore, we diluted the cells to obtain on average one infected cell per well, assuming a burst size ranging from 3 to 300 copies per cell, with at least 5 keratinocytes in the highest dilution.

Cells were serially diluted three times with a 1:100^1*/*3^ dilution factor, to obtain a range from 1 to 100, with homogenization between dilutions by pipetting up and down. Five replicates of 2µL aliquots of each cell dilution were deposited in a 96-well plate. Cells were lysed on the plate using a lysis buffer composed of Proteinase K 0.1mg/mL, Tween 20 0.2% v/v, in EDTA 1mM, Tris-HCl 10mM pH 8.2. The plate was then incubated for 4 hours at 56°C for cell lysis and 10 minutes at 95°C in a thermocycler for proteinase K inactivation.

### qPCR

To avoid any bias arising from pipetting and transfer, qPCR was performed on the same 96-well plate in which cells were lysed. After vortexing the plate, we added 11.2µL of the qPCR mix. The qPCR mix was composed in final concentrations of 5µM of forward and reverse primers targeting the cellular *albumin* gene, 4µM of forward and reverse primers targeting HPV16 *E6*, and SensiFAST^™^ Probes No-ROX Kit (MERIDIAN Bioscience: 2µM of *albumin* probe coupled with FAM, 1µM of HPV16*E6* probe coupled with Hex). qPCR reactions were performed on a LightCycler^®^ 96 (Roche Diagnostics). The cycle steps and primers sequences are detailed in the Supplementary Materials.

To create HPV standards, we used plasmids graciously provided by the international HPV reference center from the Karolinska Instituet (https://www.hpvcenter.se). Each plasmid contained a full HPV16 genome. We quantified the total DNA amount in purified plasmid preparations using Qubit to determine the total number of plasmid copies and then diluted accordingly. To create human genome standards, we used genomic female Human DNA (Promega) for the *albumin* standard, which was prepared to a known concentration of 204ng/µL.

We evaluated sensitivity and specificity of our qPCR duplex method by measuring the HPV16 *E6* and *albumin* standards in six replicates, using 1:2 serial dilution and with final concentration of *albumin* and HPV16 *E6* ranging from 1 to 64 copies per well, following [24].

This enabled us to estimate the limit of detection (lod), *i*.*e* the number of copies per well above which we can detect the presence of DNA with more than 95% certainty, and the limit of quantification (loq), *i*.*e* the number of copies per well above which the coefficient of variation is less than 35%, following [24].

### Flow cytometry

#### Sample preparation

A 1mL aliquot of the cervical smears was filtered on 70µm cell strainer, cells were pelleted via centrifugation at 600xg for 10 minutes, washed twice with PBS, and resuspended in 100µL PBS with 2% Bovine Serum Albumin (BSA).

Cells were labeled using 1:50 antibody anti pan-keratin (clone C11, Cell Signaling Technology, Danvers, MA, USA) targeting keratin 4, 5, 6, 8, 10, 13 and 18, coupled with Alexa Fluor 647 (AF-647) for 20 minutes in the dark, on ice. Finally, cells were re-suspended in 250 µL of PBS 2% BSA and stored on ice until acquisition using a flow cytometer (Novocyte, Acea).

#### Sample analysis

The resulting fcs files were processed with the Floreada analysis tool available at floreada.io (accessed on 5 April 2024). In order to exclude debris, which displayed low FSC values, from the analyses, gating in the FSC vs. SSC plot was performed for every determination. Gating in the FSC-H *vs* FSC-A plot was also performed to exclude doublets and multiplets in cell cycle analyses.

Finally, leukocytes, expected to be smaller than keratinocytes, were identified as the cells with low FSC values and confirmed by a lower AF-647 fluorescence signal. The gating was adjusted accordingly for all samples.

#### Statistical analysis

Data analysis was performed using R v.4.3.2. Joint estimation of the proportion of infected cells and the number of viral copies per infected cell was performed using a custom Bayesian statistical model with the RStan package (scripts available online at https://gitlab.in2p3.fr/ete/hpv16_copy_number).

#### Bayesian statistical model

For each sample dilution (*i*.*e*. qPCR well), the qPCR quantified the number of HPV16 genome copies (*h*) and keratinocytes (*k*). From that, we estimated a “latent” discrete variable (*i*) representing the number of infected keratinocytes in each dilution.

Then, for each sample, we jointly estimated the proportion of infected cells (*p*) and the number of viral genome copies per infected cell (*c*). For all samples, we estimated a single measurement error proportional to the log of *h*, denoted *σ*. We neglected the uncertainty in *k*.

We also estimated the effects of covariates on *p* and *c*, summarised in a matrix, *X*[*j*, 3]. Each row of *X* represents a sample; the first column is the intercept, the second is the cytological status (‘normal’ or ‘LSIL’), and the third is the proportion of leukocytes (a proxy for inflammation). We then estimated a regression coefficient vector for each parameter governing the HPV viral load, *i*.*e. β*_*p*_[*l* + 1] and *β*_*c*_[*l* + 1].

Furthermore, we assumed an unexplained variance between samples of parameters *c* and *p*, using a random effect of the sample, following a normal distribution with respective standard deviations *σ*_*c*_ and *σ*_*p*_.

#### Model likelihood

The steps leading to the full Bayesian model and its likelihood are described in the Supplementary Methods. The likelihood of the full model can be written:

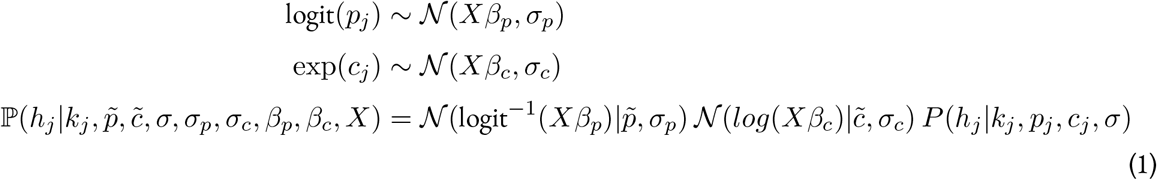

where ℙ (*h*|*k, p, c, σ*) depends on the value being above or below the detection and quantification limits of the qPCR:

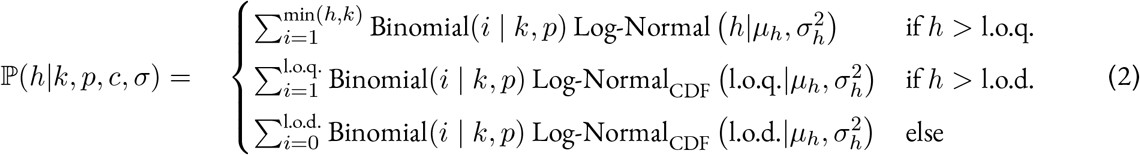

where Log-Normal_CDF_ indicates the cumulative distribution function of the *Log-Normal* distribution.

**Priors**. We assumed the following priors for the variables:

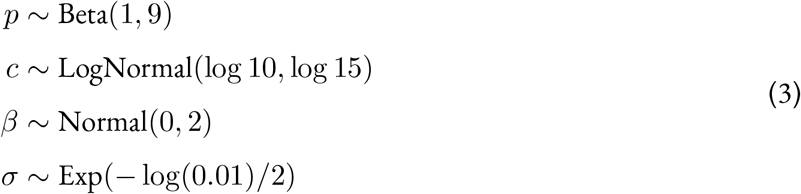

These were chosen for the following reasons. Regarding *p*, we expect a minority of the cells to be infected in the absence of cervical lesion, given that earlier results showed that smears with more than 2% of cells expressing *E6* mRNA were associated with high-grade lesions [25]. Note that this value could underestimate the total number of infected cells because some cells could be however infected without *E6* expression. Therefore, we assumed a prior with a mode centered on 0, but a wide variance and a mean at 0.1. Regarding *c*, given the large degree of uncertainty, we opted for a poorly informative prior with 95% of the values between 1.6 and 618 (median of 10 copies and standard deviation of 200). We kept as well a poorly informative prior for the regression parameters*β* for lesion status and inflammation.

## Results

### Statistical model validation

We assessed the potential biases of our statistical model using simulated data with known parameters, by comparing the mean estimate obtained from 200 simulations for each set of parameters, using systematically the same priors.

Existing statistical methods to infer the proportion of infected cells in a sample following limiting dilutions typically ignore both the mean number of viral copies per infected cell and the limit of detection of the assay [26, 27].

The median estimates of the mean burst size and the proportion of infected cells were close to the simulated parameters (Fig. 1). It started to diverge more from the actual value when the mean burst size was above 50. As expected, the estimation also became worse when the mean number of HPV copies per cell was below the limit of quantification (assumed to be 8 HPV copies).

**Figure 1.**
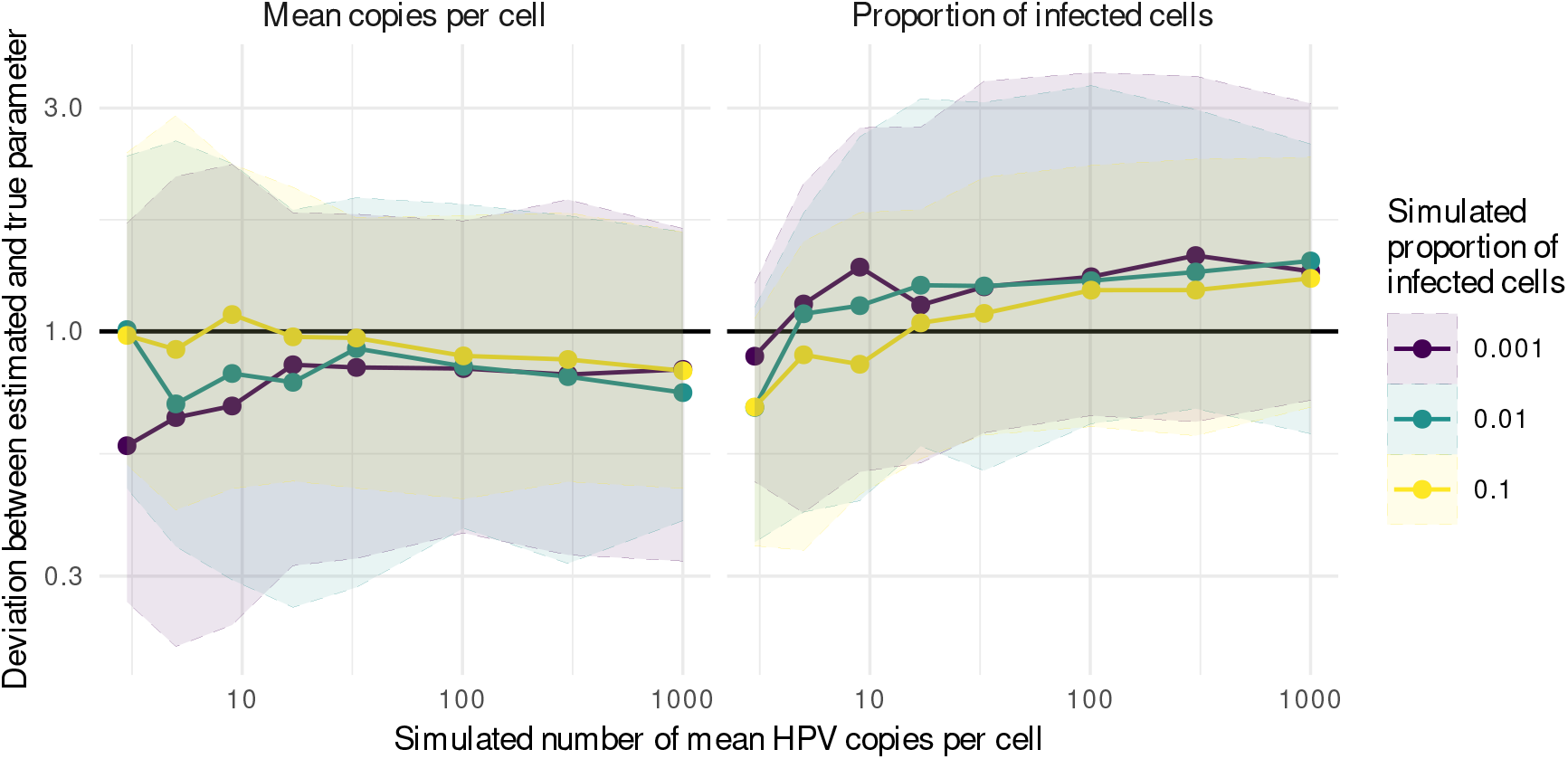
Parameters estimates of simulated data, as a function of the target number of mean HPV copies per cell. The colors indicate the simulated proportion of infected cells. For each set of parameters, we performed 200 simulations, assuming stochasticity in the dilution process in addition to the uncertainty of the qPCR assay. We then recovered the median estimate for the mean copies per cell and the proportion of infected cells. Shaded areas show the 95% quantile of the median estimates and central thick lines present the median estimate value.

Within the optimal range of burst size of 8 to 300 copies, our method tended to underestimate the mean burst size by 15% on average. Conversely, it tended to overestimate the proportion of infected cells by 15% on average (Fig. 1). However, even outside this range, our estimates remained in the order of magnitude of the true value, and the expected value was in the 95% credible interval (CrI) in 80 to 100 % of the simulations (Fig. S6).

The true distribution of the simulated number of HPV copies is the sum of the number of copies per infected cells, (which follows a zero-truncated Poisson distribution). However, it was approximated by a lognormal distribution on the statistical model. This can explain the observed bias.

### Estimates from SiHa cells

We then used a mixture of SiHa cells and HPV-negative cervical swabs in known proportion, as a proof of concept for our method.

We experimentally estimated an average of 1.7 copies of HPV16 genome copies per copy of albumin in these cells (95% confidence interval (CI): 0.7 - 4.7 copies). This represents on average 3.4 copies of HPV16 per cell when considering 2 copies of albumin per cell. These estimates are largely consistent with our experimental system, as SiHa cells contain on average 1 to 3 integrated HPV16 genome copies per cell [28]. Although the SiHa genome is nearly triploid, it only contains two copies of albumin gene per cell, since their chromosome 4, which harbors this gene, is present in two copies [29].

We mixed a known number of HPV-negative cervical keratinocytes with 1%, 10%, or 25% of SiHa cells. We then proceeded using our limiting dilution protocol to estimate the proportion of infected cells and the “burst size”.

We found that the expected values always fell within the 95% CrI of our estimates (Fig. 2), except for the number of HPV16 copies per infected cell when the proportion of infected cells is 1%. Our Bayesian statistical model tends to over-estimate *c* and to under-estimate *p* (Fig. 2), especially when the frequency of infected cells (*p*)is low, which is consistent with the numerical simulations when the number of viral copies is very low (Fig. 1).

**Figure 2.**
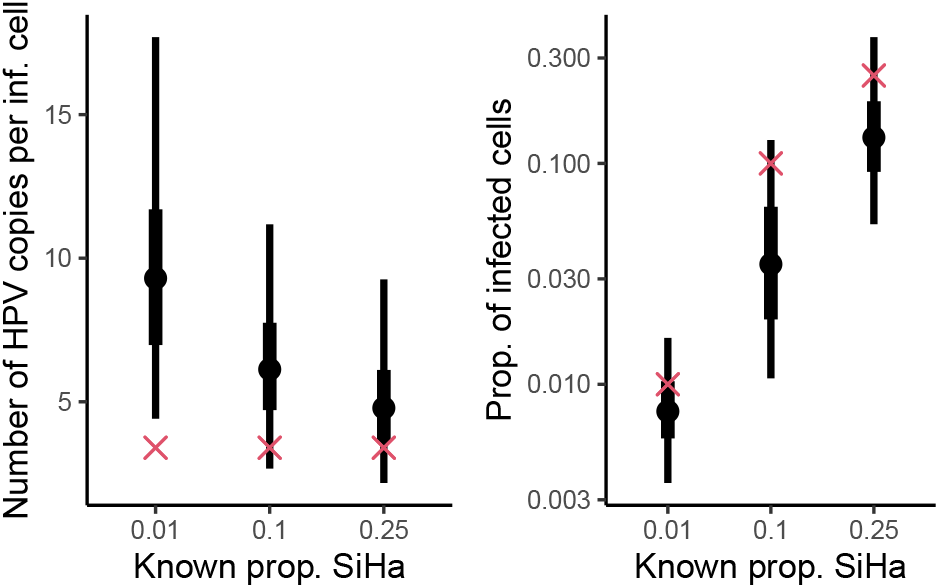
Posterior distribution for the estimated mean number of HPV copies per infected cell and the proportion of infected cells from cell cultures. Results are the aggregated value of two technical replicates. The red cross indicates the expected value, as per experimental design. The dot represents the median posterior value, the thick line shows the interquartile range, and the thin line, 95% CrI.

### Estimates from cervical smear clinical samples

We then applied the limiting dilution protocol, combined to our Bayesian model tested above to jointly estimate the proportion of infected cells (*p*) and the mean number of HPV copies per infected cell (*c*) of cervical smears. These samples were originating from patients infected by HPV16 and no other high-risk genotype. There were *n* = 26 patients without any lesion and *n* = 12 with an LSIL.

First, to verify the accuracy of our estimates of leukocytes proportion made with microscope counts, we randomly selected 20 samples to analyse them with flow cytopmetry. The two measurements showed indeed a good agreement (Pearson correlation coefficient *ρ* = 0.65 (95% CI: 0.28-0.85), S5).

There was an important variability in viral load among samples (Table 1). However, when estimating the impact of both lesion status and proportion of inflammatory cells on the viral load in the same model, viral load tended to be higher in samples with an LSIL (mean log10 HPV copies/cell difference of 1.0 with F(35,1) = 8.1 and p = 0.09). This is consistent with other cross-sectional studies showing that viral load is weakly associated with cytological status. Conversely, viral load was not affected by the presence of inflammatory cells (F(35,1) = 4.5 and p = 0.21).

**Table 1:** Summary of the 38 samples analysed stratified by cytology results.

|  | Normal cytology<br>mean (IQR) / n (%) | LSIL<br>mean (IQR) / n (%) |
| --- | --- | --- |
| Number of samples | 26 (68%) | 12 (32%) |
| Viral load (copies per cell) | 0.14 (0.02-0.83) | 1.01 (0.10-14.3) |
| Presence of inflammation (on the anapathological report) | 10 (38%) | 8 (67%) |
| Proportion of leukocytes (microscope counts) | 52% (34-69) | 67% (56-79) |

#### An important variability of the parameters between the samples

We observed an important heterogeneity in the proportion of infected cells between samples, with a median estimate ranging from 0.004% to 95% in the 38 samples (Fig. 3A). The mean number of HPV copies per cell varied in a lower range, with median estimates ranging from 3 to 116 (Fig. 3A). Importantly, in 6 samples out of the 12 with LSIL we always obtained infected cells, while this was not the case in any of the samples with normal cytology, even when diluting up to less than 5 keratinocytes per well (Fig. S8). This resulted in a higher estimate of the proportion of infected cells in those samples for which we could not generate HPV-negative dilutions, compared to the other samples (Fig. 3A).

**Figure 3.**
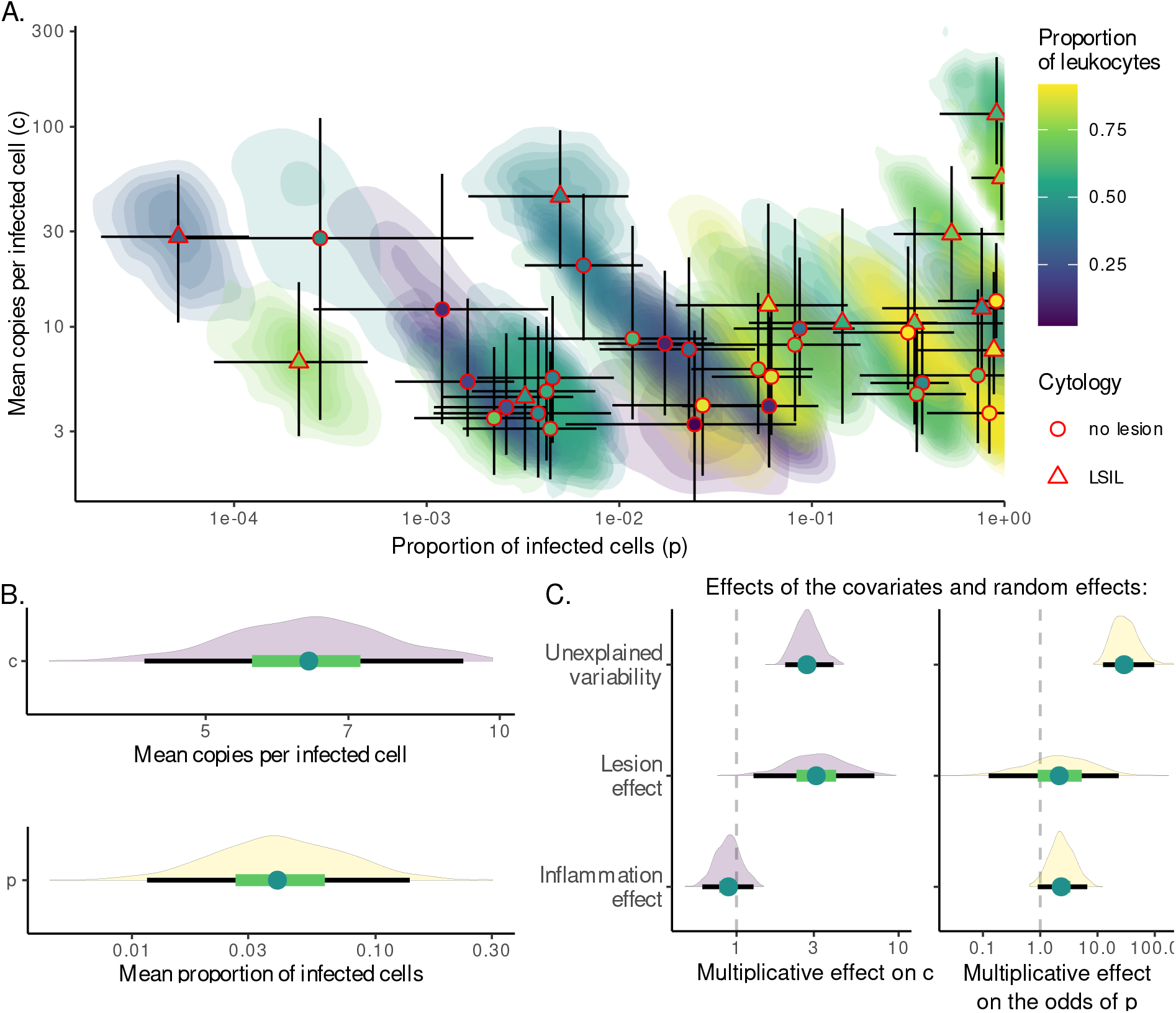
Estimating the proportion of infected cells and the number of virus copies per infected cells from cervical smears. A. Mean number of HPV copies per infected cell (*c*) as a function of the proportion of infected cells (*p*). Each point or triangle represents the median estimate of one sample. Colors indicate the mean proportion of leukocytes in the sample, which is a proxy for local inflammation level. The posterior density is represented by the color shades, which illustrates the covariation of the two estimates. The lines in both directions represent 95% CrI of each parameter. B. Model parameters values. The reference was a sample with normal cytology and average inflammation status. C. Effects of two covariates, and the unexplained variability between samples on parameters *c* and *p*. The effects on *c*, which is log-tranformed, must be interpreted as a multiplicative effect. The effects on *p*, which is logit-transformed, correspond to a multiplicative effect of the odds. For the lesion status, the reference is the normal cytology, and for the proportion of leukocytes in the sample, we used a (scaled) linear effect. The unexplained variability represents the standard deviation of the sample random effect associated with *p* and *c*. The dots represent median values, the thick green lines the 50% CrI, and the thin black lines the 95% CrI.

As anticipated, we observed a good correlation between the viral load measured in the bulk sample and the product of our estimates for the proportion of infected cells and the burst size (*p* × *c*), adjusted for the proportion of keratinocytes (Pearson’s correlation coefficient for the log viral load: 0.89, with a 95% CI from 0.79 to 0.95). Lower viral loads exhibited greater variation. Additionally, our estimates from the limiting dilution assay tended to underestimate the viral load compared to the bulk estimate (Fig. S9).

### The lesion, but not inflammation level, partly explain the variability around the mean parameters

With our model, we could estimate an average value for the proportion of infected cell and the mean number of viral copy per infected cell, as well as an effect of the inflammation and the proportion of leukocytes in the sample.

The average estimate, in a sample without lesion, was that 4% of the cells in the samples were infected, with a 95% CrI of 1% to 15% (Fig. 3B). There was no correlation between the fraction of infected cells and the presence of a lesion. Consequently, a significant degree of unexplained variability remained between samples (Fig. 3C), which was considerably higher than the effect of the covariates.

The estimated average number of viral genome copies per cell in a sample without cytological lesion was found to be 6, with a 95% CrI of 4 to 9 copies (Fig. 3B). Notably, the number of viral genome copies per cell was found to be significantly higher in the presence of LSIL, averaging 17 copies. This represented a threefold increase (95% CrI: 1 to 7 times) compared to samples without cytological lesions (Fig. 3C). Furthermore, the unexplained variability between samples in burst size was of a similar magnitude to the effect of the cytological lesion.

## Discussion

Despite its public health importance, the details on the natural history of HPV16 genital infection, and, in particular, its quantitative aspects such as the frequency of infected cells and the mean number of viral copies per infected cell, are largely unknown. We developed a Bayesian statistical model able to jointly estimate the proportion of infected cells and the mean number of copies per cell, while taking into account the sensitivity of the assay to analyse a simple and robust limiting dilution approach.

We validated our protocol using numerical simulations and cell culture-based experimental data. Our statistical model accurately recovered true values most of the time, with a slight systematic bias towards overestimating the fraction of infected cells and underestimating the viral genome load per cell due to unavoidable statistical approximations. Data from controlled cell cultures confirmed that our model correctly estimated the number of viral copies per cell, even below the assay’s limit of quantification. Additionally, the supplementary step of mixing HPV-negative samples with SiHa cells might have introduced more variability compared to estimates from infected cervical smears.

We estimated that the number of HPV copies per infected cell in samples with normal cytology or LSIL ranges from 3 to 116 copies, which is slightly lower than previous findings in high-grade lesions [18, 19]. These estimates are also lower than those obtained experimentally from lesions in the Rabbit Oral Papillomavirus (ROPV) model [17]. Although the keratinocyte origin in our samples was not further characterized, we can infer that the brush collects cells from the parabasal layer, which contained 10^3^ to 10^4^ PV copies per cell in the ROPV model. This discrepancy can be attributed to the highly productive nature of ROPV lesions in the epithelium, which differ significantly from HPV16 cervical infections without lesions.

The estimated proportion of infected keratinocytes varied widely, typically ranging from one in 20,000 to nine in ten. Despite this variability, the high proportion of infected cells ensures a good probability of detection given the typical cell count in a cervical smear, which is considered satisfactory with at least 8,000 to 12,000 well-preserved cells according to The Bethesda system [23]. If only a few cells contained viral genomes, stochastic processes might have a greater influence on viral load dynamics. However, HPV infection kinetics in young women show a stable viral genome load plateau lasting several months in young asymptomatically infected individuals [30], or a steady decrease in viral load in the general population [31, 8]. The significant proportion of infected cells does not exclude the role of stochasticity in other stages of the virus life cycle, particularly in the basal layers of the epithelium [32, 33].

We included two explanatory variables in our statistical model: the natural history of the infection and the local inflammatory response. To test the hypothesis that low-grade lesions are associated with high viral genome load, reflecting either increased burst size or frequency of infected cells, we found that the viral genome load per infected cell in LSIL samples was nearly three times higher than in samples without lesions, although the proportion of infected cells remained unchanged. This aligns with studies showing higher viral loads in LSIL, especially HPV16-positive LSIL [10, 12]. This finding is consistent with the notion that E4 protein expression, which is high in productive upper epithelial layer [34] and enhances genome amplification and virus synthesis [35], increases with LSIL appearance but decreases in high-grade lesions [36].

The proportion of leukocytes showed a weak positive correlation with the proportion of infected cells, although this was not significant at the 5% false-positive threshold. This may indicate that a higher number of infected cells induces more inflammation in the cervix. However, this observation could also be influenced by the sensitivity of our estimates based on keratinocyte counts from microscopy (Fig. 3C). Overestimating leukocytes would result in an underestimation of keratinocytes, leading to an overestimation of the infected cell proportion and an underestimation of viral copies per infected cell.

We observed important heterogeneity among the twelve LSIL samples, with six consistently showing positive HPV16 qPCR results across serial dilutions. This variation may reflect different infection dynamics, where most LSIL cases regress, but some progress to high-grade lesions. We hypothesize that the six samples with consistently positive HPV16 qPCR, indicating a high proportion of infected cells, could be at higher risk of progression to high-grade lesions, while the others may regress. Progressing cases are typically associated with increased expression of cell proliferation markers, such as cellular p16 protein [36] and viral E6 or E7 proteins [37]. This association between LSIL with more infected cells and higher progression risk aligns with Baumann *et al*. [6], who reported that LSIL with higher viral load significantly increased the risk of lesion progression.

Overall, the important variability in the proportion of infected cells observed in our samples was poorly accounted for by our covariates. This is evidenced by the substantial standard deviation of the associated sample random effect, indicating that a majority of the variability in the proportion of infected cells between samples remains unexplained. Further investigations are needed to elucidate the proportion of infected cells in a cervical smear. The sampling process itself, conducted by different gynecologists, may have introduced additional variability, potentially due to inconsistent swabbing of the cervix. Cervical HPV infections are typically confined to small areas of the cervix, as observed in women with infections but no lesions [38]. Additionally, the immune response likely influences the frequency of infected cells, impacting the dynamics of asymptomatic infections [30], particularly in conjunction with the vaginal microbiota [39]. Future studies incorporating cell proliferation and immune response markers could be instrumental in determining the biological mechanisms underlying this heterogeneity.

Having better quantitative estimates of the HPV infection life cycle could inform mechanistic models. In this perspective, the number of viral genome copies per cell is a key parameter, but it remains largely poorly explored [40]. In particular, very few studies have modeled the different steps of the HPV natural history in a mechanistic way [33]. To our knowledge, only [41] include the appearance of pre-cancerous cells during the infection, assuming that they have a higher carrying capacity and that they do not differentiate, thus leading to something more similar to a high-grade lesion. The observation here reported that the burst size increases in the LSIL samples but that the proportion of infected cells does not increase could guide the construction of models simulating the risk of appearance of a lesion.

Our combination of statistical and molecular approaches could be particularly useful to better understand the biology of the HPV infection when there is no high-grade lesion, as it is very often not possible to gather observations from biopsies in the case of low-grade lesions. Furthermore, our method relies on cervical smears obtained from cervical cancer screening, which are broadly available.

One limitation of our method is the use of samples in Thinprep medium, a methanol-based fixative that likely permeabilizes cells, potentially resulting in virion or in viral genome loss. Additionally, highly productive cells may degrade during handling, releasing viral DNA and/or virions into the supernatant, thereby creating a background of positive wells with low copy numbers. This could account for the observed lower mean number of HPV copies per cell in high viral load samples compared to bulk estimates (Fig. S9). Consequently, the proportion of infected cells may be overestimated, explaining why six LSIL samples did not yield any HPV-negative dilutions, even with low cell counts. Another potential bias could relate to the cell filtration step, which might preferentially remove aggregated keratinocytes that stem from the upper epithelial layers and that likely contain most viral genome copies, thus increasing the proportion of smaller leukocytes in some samples. Finally, our method requires an optimized qPCR assay with low detection and quantification limits to minimize information loss when the number of infected cells per well is near one and burst size is low. Future refinement of our estimates could be achieved using more accurate methods, such as digital PCR, which offers better quantification limits.

Our work focused on HPV16 single infections, to avoid confounding factors, and because this genotype is the most oncogenic [2]. Our method could be easily implemented in other contexts to increase the amount of data. In particular, this could give useful insights into the important difference in risk of oncogenicity between HPV genotypes.

## Data Availability

All data produced are available online at https://gitlab.in2p3.fr/ete/hpv16_copy_number

## Acknowledgements

We thank the CNRS, the IRD, and the IRD itrop HPC (South Green Platform) at IRD Montpellier for providing HPC resources that have contributed to the research results reported within this paper. We also thank the ETE modelling team for discussion.

## S1 Ethics

This study was conducted in accordance with the tenets of the Declaration of Helsinki. In accordance with French legislation (Public Health Code, as amended by Law No. 2004-806 of 9 August 2004, and Huriet-Sérusclat Law 88-1138 of 20 December 1988), and as this study only involved data extracted from medical records, informed consent of the patients was not required.

HPV16-positive samples used were stored in a biobank for which a declaration on the preparation and storage of human samples for research purposes has been submitted to the Ministe`re de l’Enseignement Supérieur et de la Recherche (n°DC-2014-2086).

HPV-negative samples came from the study “Health in climacteric women belonging to NGOs. PAPI-LONG study”, validated by the Ethical committee of the CEI DGSP-CSISP (Valencia, Spain), with number 20210604/10/01.

## S2 Supplementary methods

### S2.1 qPCR

**Table S1:** qPCR cycle description.

| Temperature (°C) | Duration (sec) | Cycles |
| --- | --- | --- |
| 95 | 300 | 1x |
| 95 | 10 | 40x |
| 60 | 30 |  |

**Table S2:**
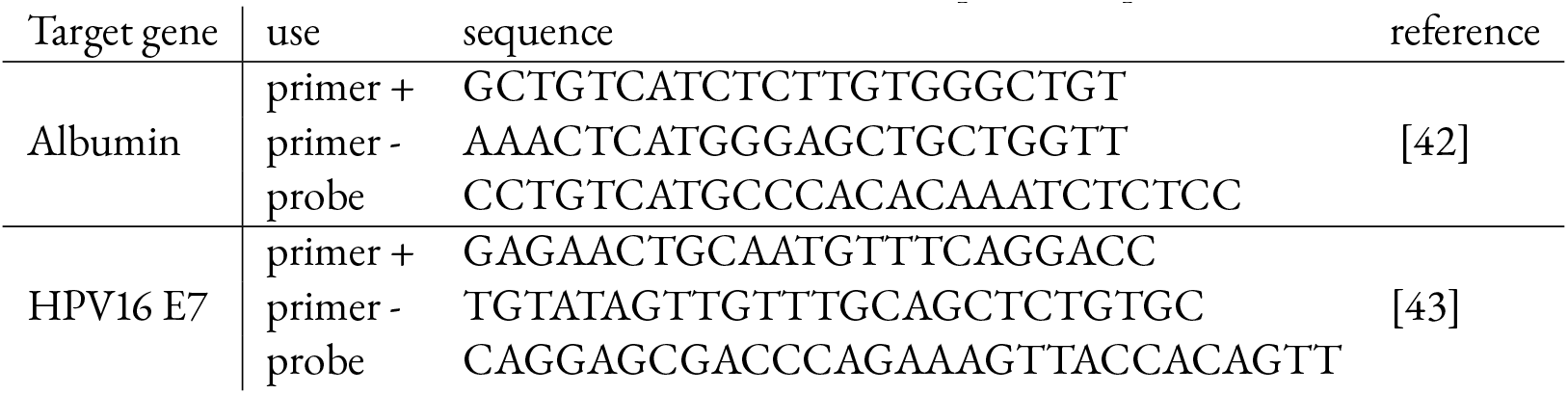
Primers and probes sequences.

### S2.2 Bayesian model likelihood

For each sample, we measured the number of HPV16 genome copies (*h*) and of keratinocytes (*k*). We jointly estimated the proportion of infected cells (*p*) and the number of viral genome copies per infected cell (*c*), using a latent discrete variable (*i*) representing the number of infected keratinocytes, and a measurement error *σ*. We simplified the problem by assuming that the uncertainty only originates from the estimate of *h*.

In the following, we first introduce our model assuming only one sample. We then extend it to several samples having the same mean. Finally, we add a potential effect of covariates into the model.

#### Model assuming only one sample

We want to estimate the likelihood of observing *h* HPV copies in a sample, given the estimated parameters, *i*.*e*. ℙ (*h*|*k, p, c, σ*). For that, we need to use an unknown parameter, the number of infected cells *i*. We can decompose the likelihood as a function of this “latent variable” *i*:

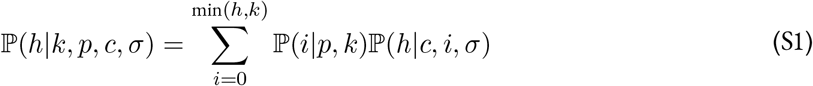

Assuming that the probability of sampling is the same regardless of the infection status, the number of infected cells can be assumed to follow a Binomial distribution.

The observed number of HPV genome copies is then the product of two sources of uncertainties: the stochasticity in the total number of copies, which we assume to be close to a Poisson process, and a measurement error *σ* that we estimate. As a consequence, we assume that the total number of HPV copies follows a log-normal distribution, with mean *c* × *i*, and variance equal to the mean. With a simple algebra computation to translate this for log-normal location and scale, this results in the distribution: 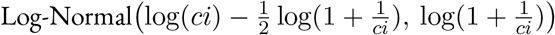. The measurement is then assumed to have an error proportional to its logged value, with mean 0 and scale *σ*, which means: 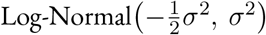. Using the convenient multiplicative property of the log-normal distribution, one simply has to sum the location and scale parameters. The resulting observed *h* is assumed to follow a log-normal distribution with location 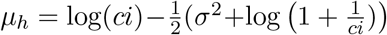, and scale 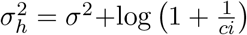 The resulting density can be estimated using a log-normal distribution. Building on equation S1, this leads to the following equation:

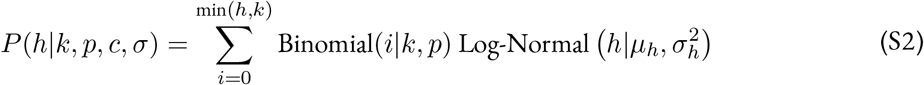

Finally, we further distinguish between the values that are above the limit of quantification (denoted ‘l.o.q.’), above the limit of detection (denoted ‘l.o.d.’), and below. This corresponded to the following likelihood:

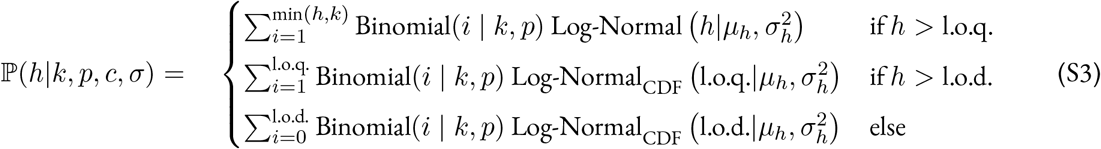

#### Model assuming several samples

We then consider several samples numbered *j* ∈ {1, …, *J*}, with the same mean proportion of infected cells 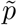 and number of copies per cell 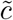. In what follows, the subscript _*j*_ denotes a parameter value estimated for sample *j*. The estimated between-samples variance associated with these variables are denoted *σ*_*p*_ and *σ*_*c*_ respectively. Our model likelihood function can then be expressed as:

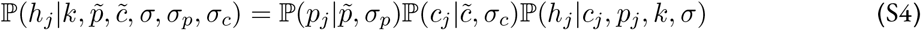

Given the nature of the data, we assume an exponential link for the error of *c* and a logit link for the error of *p*. This leads to the equations:

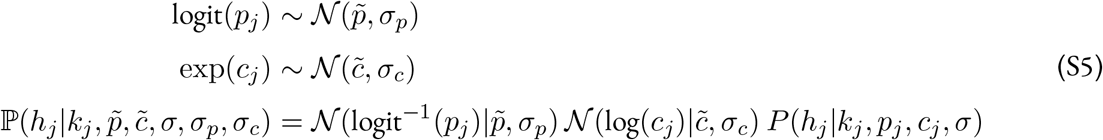

where *P* (*h*|*k, p, c, σ*) is the formula described in equation S3 and N (*m, sd*) is a normal distribution with mean *m* and standard deviation *sd*.

#### Model assuming several samples with different means

Finally, we assume that *p* and *c* can vary and be affected by *l* covariates. These are summarised in a matrix, *X*(*j, l* + 1), in which each row represents a sample, the first column is the intercept, and each of the following columns represents the value of each covariate. We then estimate a regression coefficient vector for each parameter governing the HPV viral load, *i*.*e. β*_*p*_[*l* + 1] and *β*_*c*_[*l* + 1].

The full model is now:

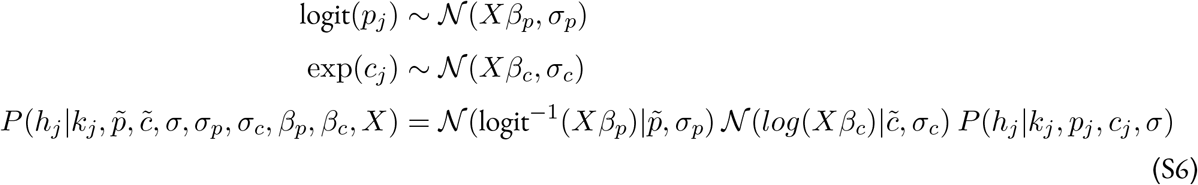

where *P* (*h*|*k, p, c, σ*) corresponds to the formula described in equation S3.

## S3 Supplementary figures

### S3.1 qPCR sensitivity

As detailed in the methods, we performed qPCR estimations with known number of HPV and albumin copies in order to determine the lod and loq.

**Figure S1:**
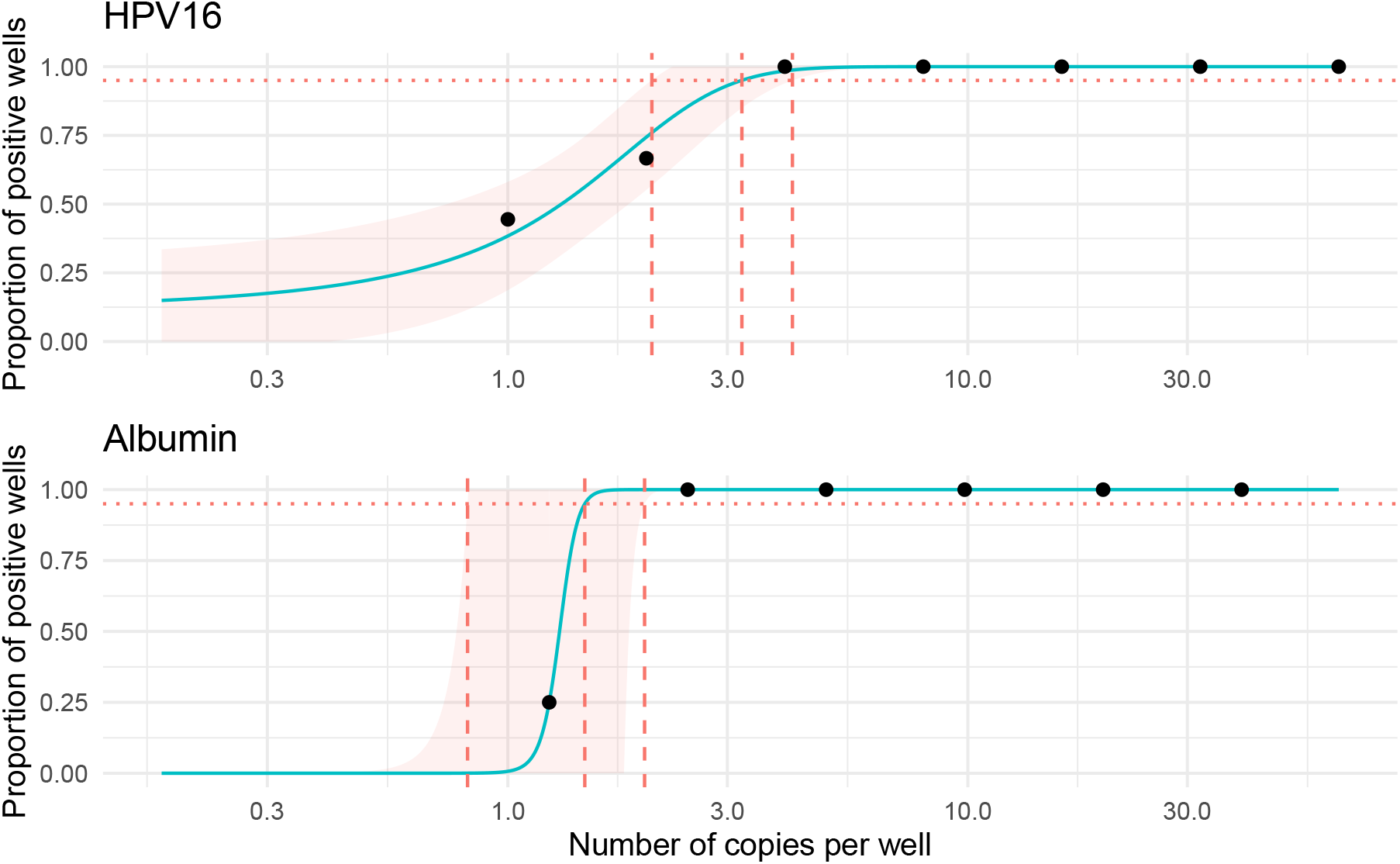
Determination of the limit of detection of the viral *E7* and the human *albumin* genes. The dots indicate the observed proportion of wells in which we detected the presence of the corresponding target as a function of the known number of copies per well. The blue line represents the maximum likelihood of a logistic fit and the red area, the 95% CI. Dashed vertical lines represent the estimate of the lod and its 95% CI.

**Figure S2:**
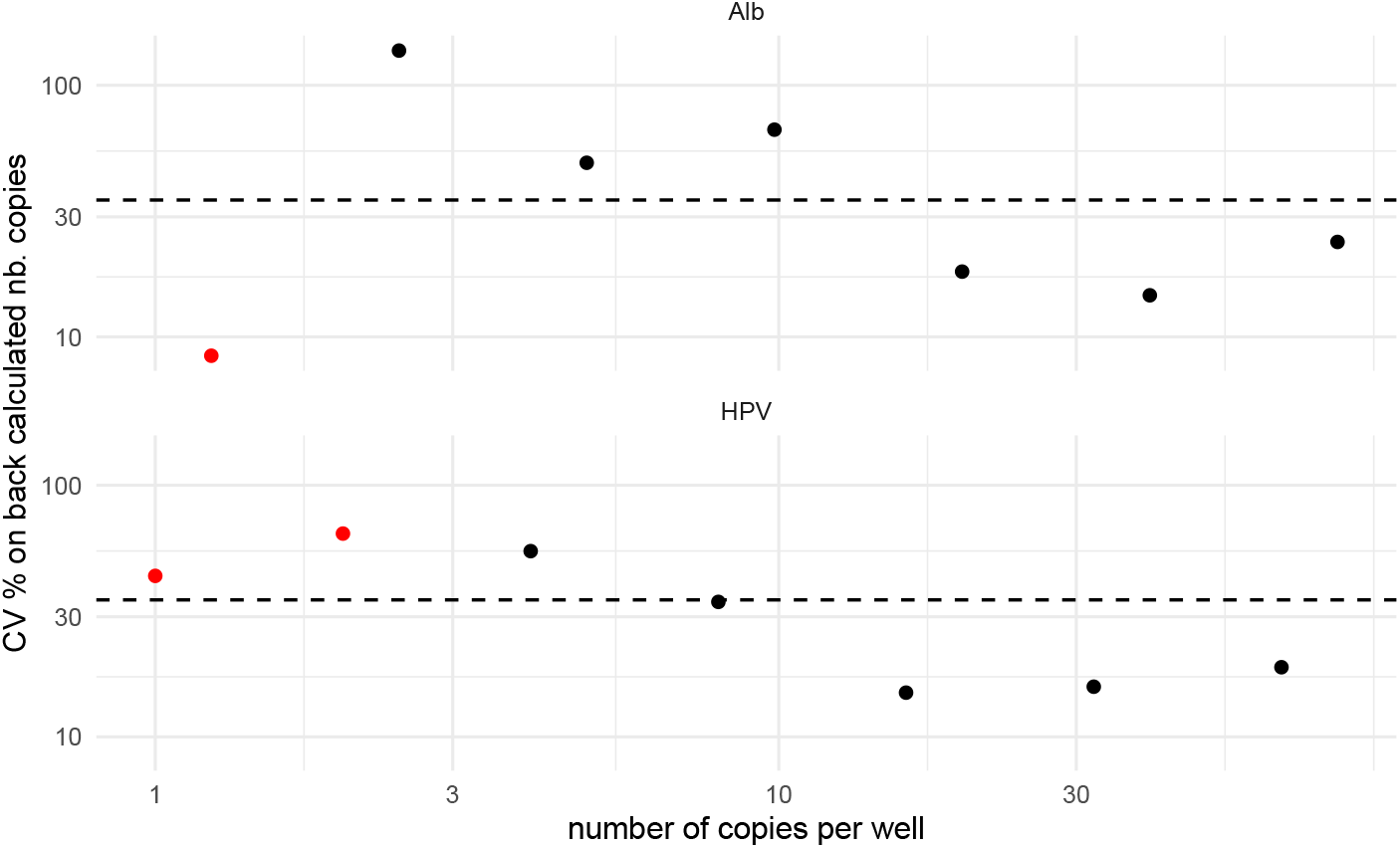
Determination of the limit of quantification of the viral *E7* and the human *albumin* genes. Coefficient of variation (CV% = 100 × SD/mean) of the back-calculated concentrations as a function of the known number of copies per well. The horizontal dashed line represents the threshold of 35%. Red dots indicate dilutions where some of the wells yielded qPCR-negative results. Those were ignored when calculating SD, resulting in a bias toward lower SD values.

### S3.2 Microscope and FCM

**Figure S3:**
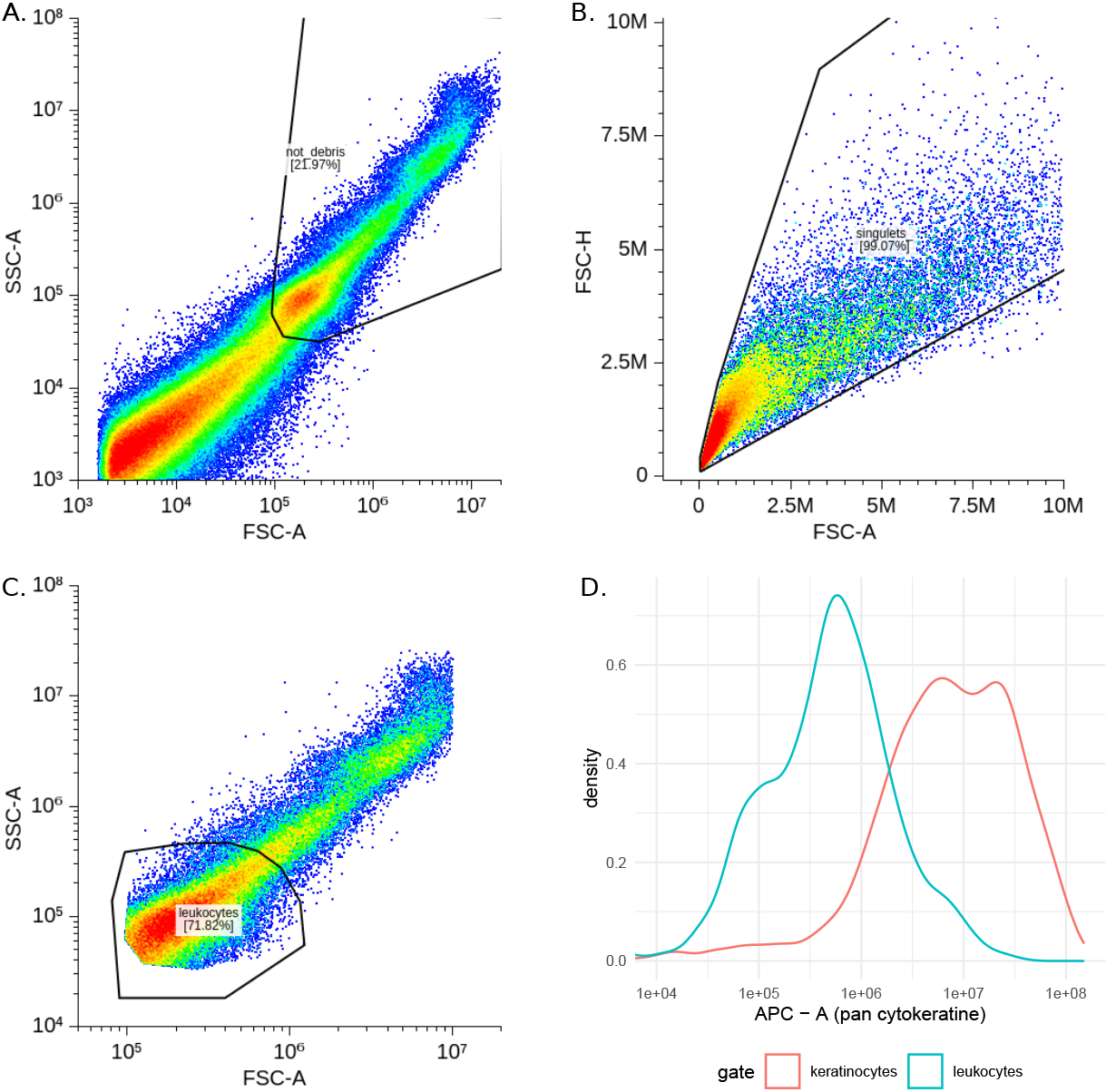
Gating steps used for the FCM analysis, on a representative sample. All 20 samples were analyzed using the same gating strategy. A. We first removed the debris based on the FSC-A/SSC-A representation on the log scale. B. We then removed doublets based on the FSC-A/FSC-H representation on the natural scale. C. We then identified the leukocytes, which are smaller than the keratinocytes. D. We confirmed that the leukocytes had a less intense signal than the other cells using an APC-pankeratinocyte antibody.

**Figure S4:**
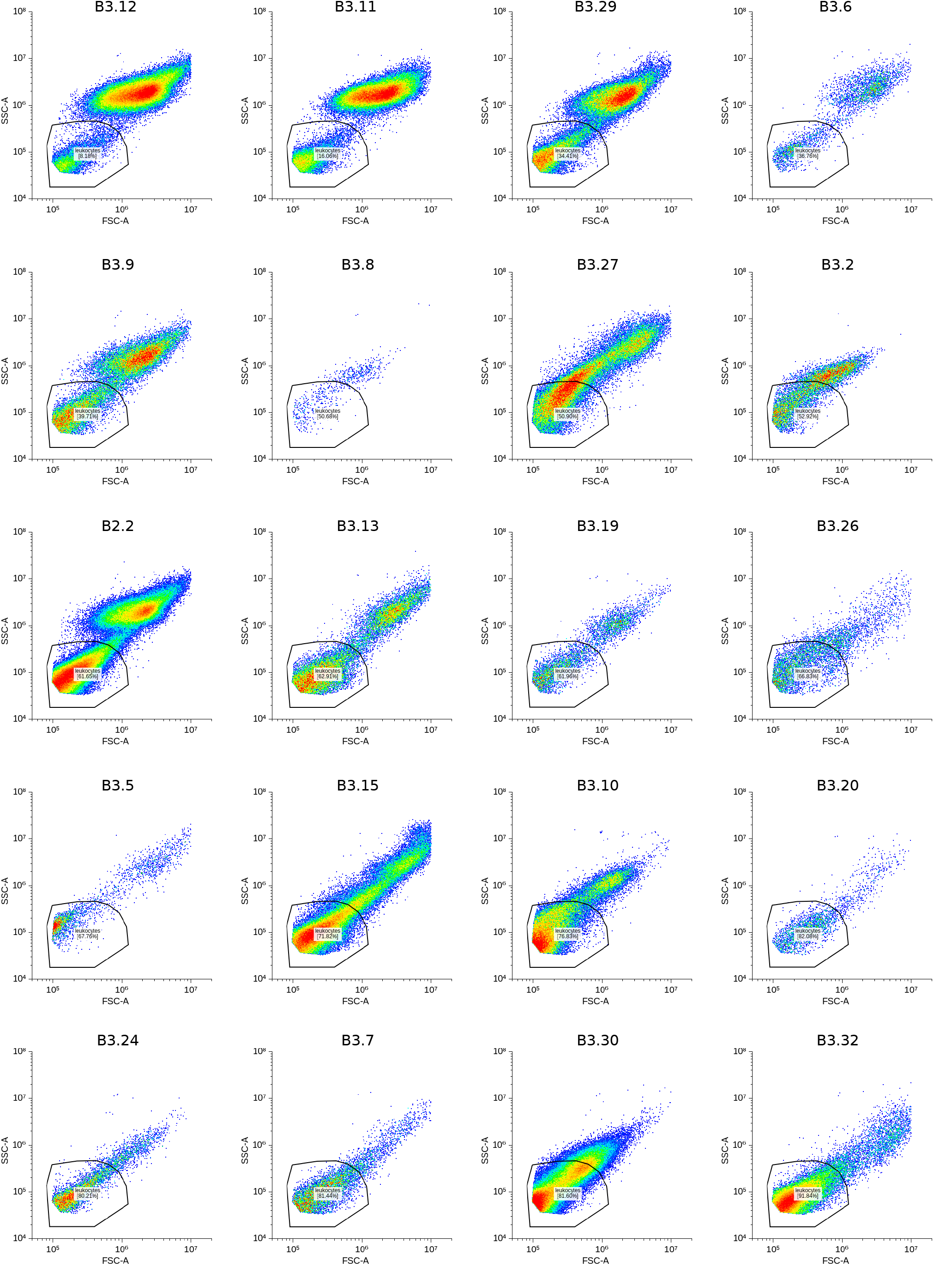
Gating of the leukocyte population on the 20 samples. All 20 samples were analyzed using the same gating strategy presented in Fig. S3.

**Figure S5:**
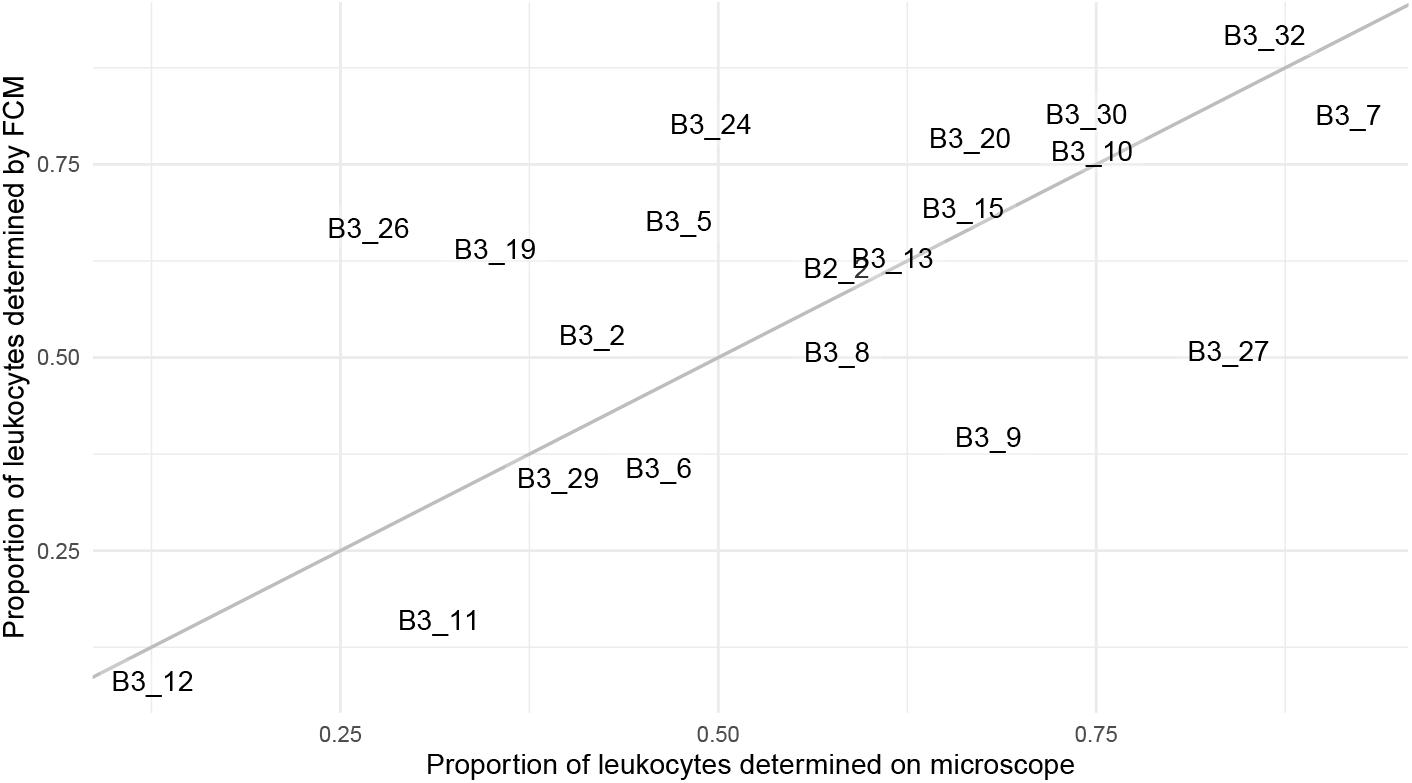
Proportion of leukocytes based on microscope counts versus on FCM analysis. In this subset of 20 samples where FCM analysis could be done, detailed in figure S4, the proportion of leukocytes had a Pearson correlation coefficient *ρ* = 0.65 (95% CI: 0.28-0.85) with the proportion of leukocytes counted in the microscope.

### S3.3 Proofs of concepts: simulations and cell culture

**Figure S6:**
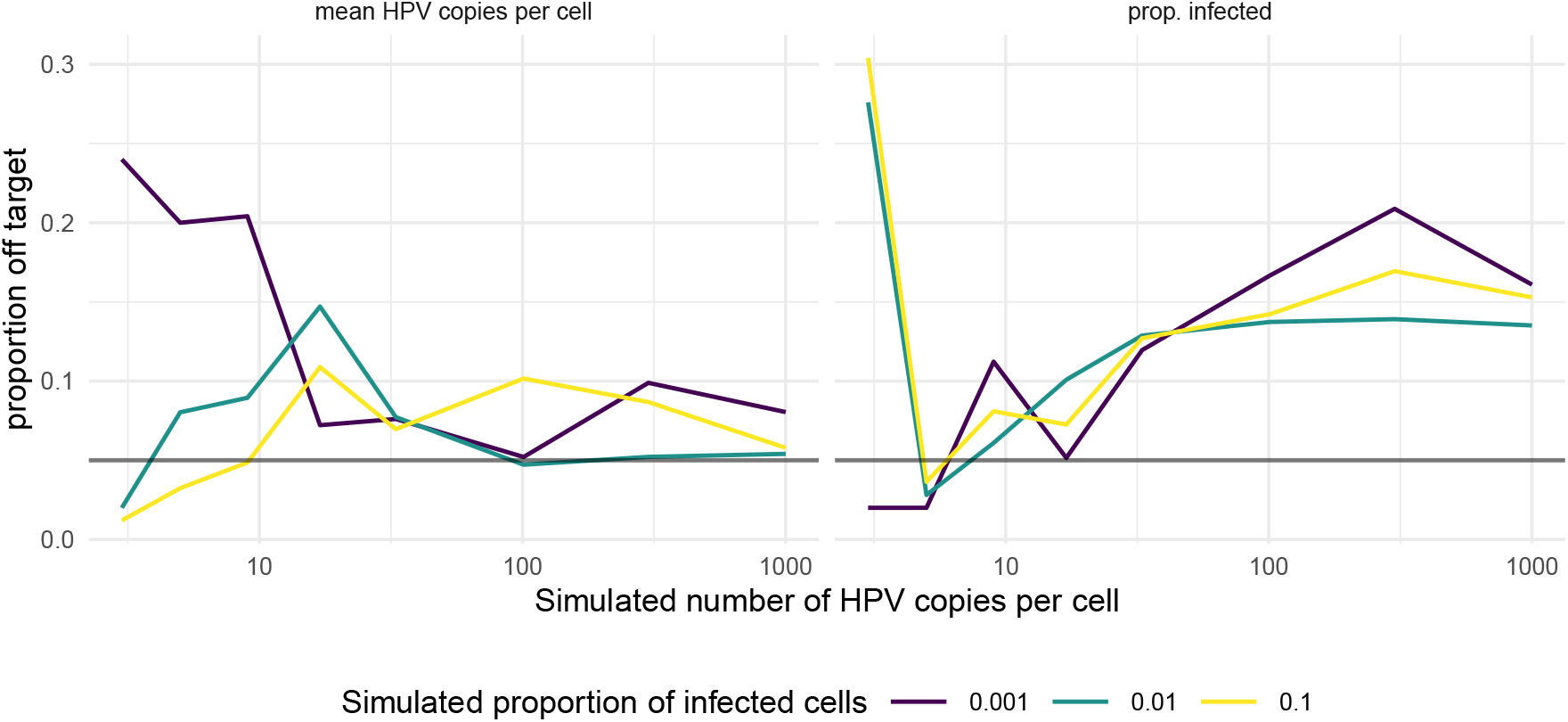
Proportion of the simulations where the 95% CrI does not include the true value. The colors indicate the simulated proportion of infected cells. For each set of parameters, we run 200 simulations, assuming stochasticity in the dilution process and uncertainty in the qPCR assay.

**Figure S7:**
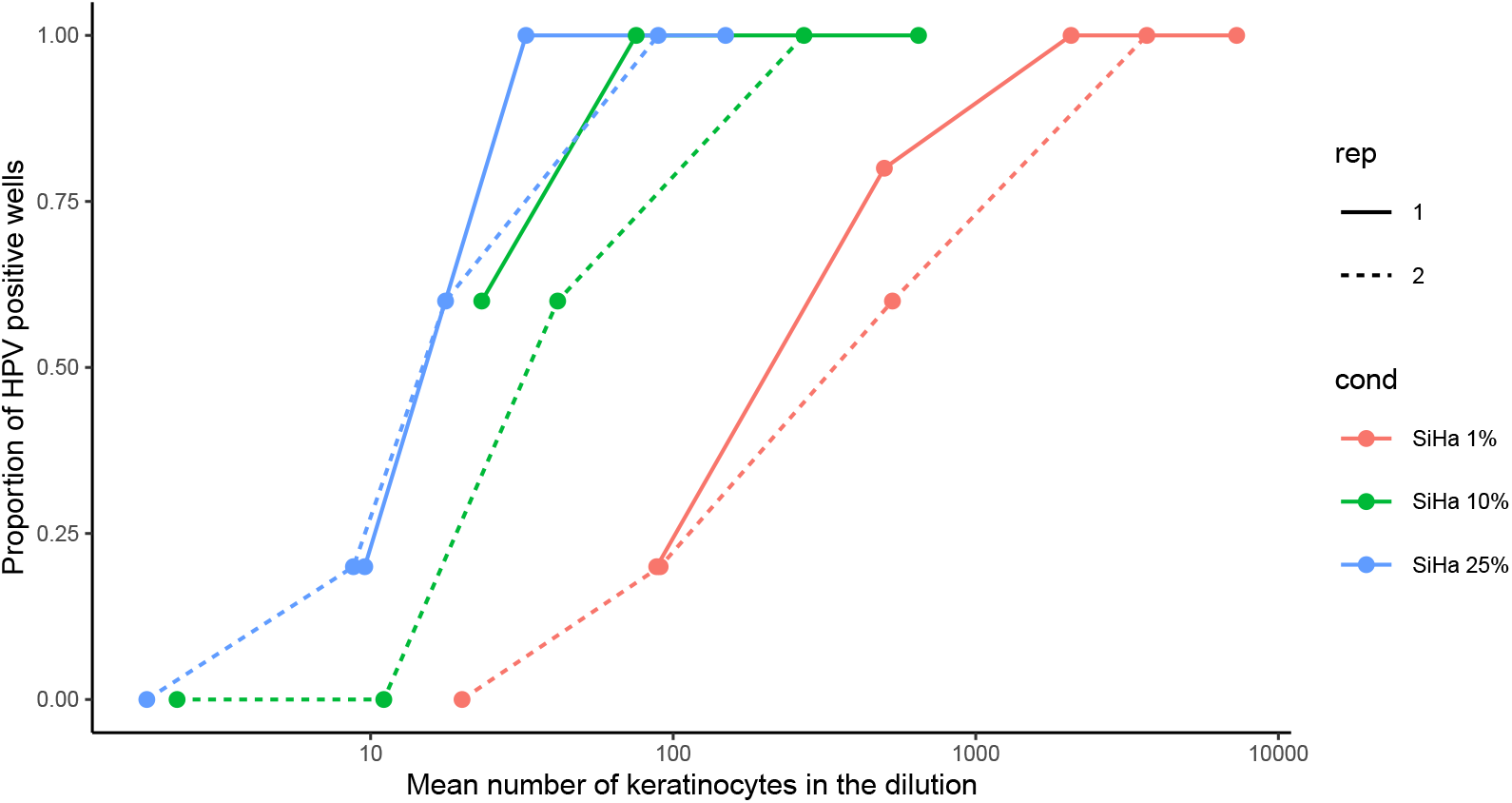
Proportion of positive wells for each dilution of the limiting dilution assay with cell cultures. The colors indicate the proportion of HPV-positive SiHa cells in the sample. The dotted lines represent a technical replicate with the same conditions.

### S3.4 Cervical samples

**Figure S8:**
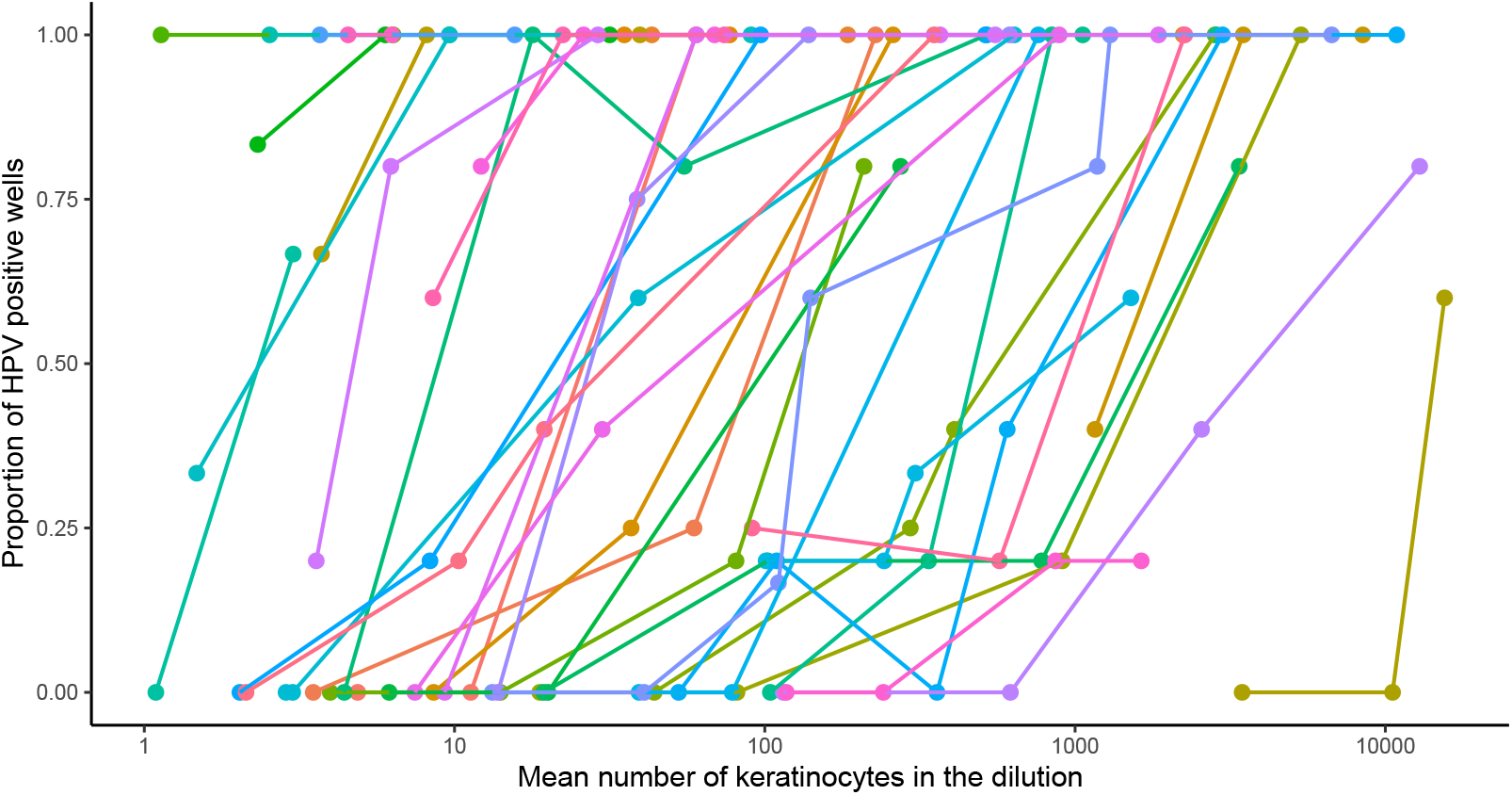
Proportion of positive wells for each dilution of the limiting dilution assay with clinical samples. Each color represents a sample. For each sample, each dilution was replicated 5 times, and we measured the frequency of the qPCR wells that yielded results above the limit of detection (4 copies).

**Figure S9:**
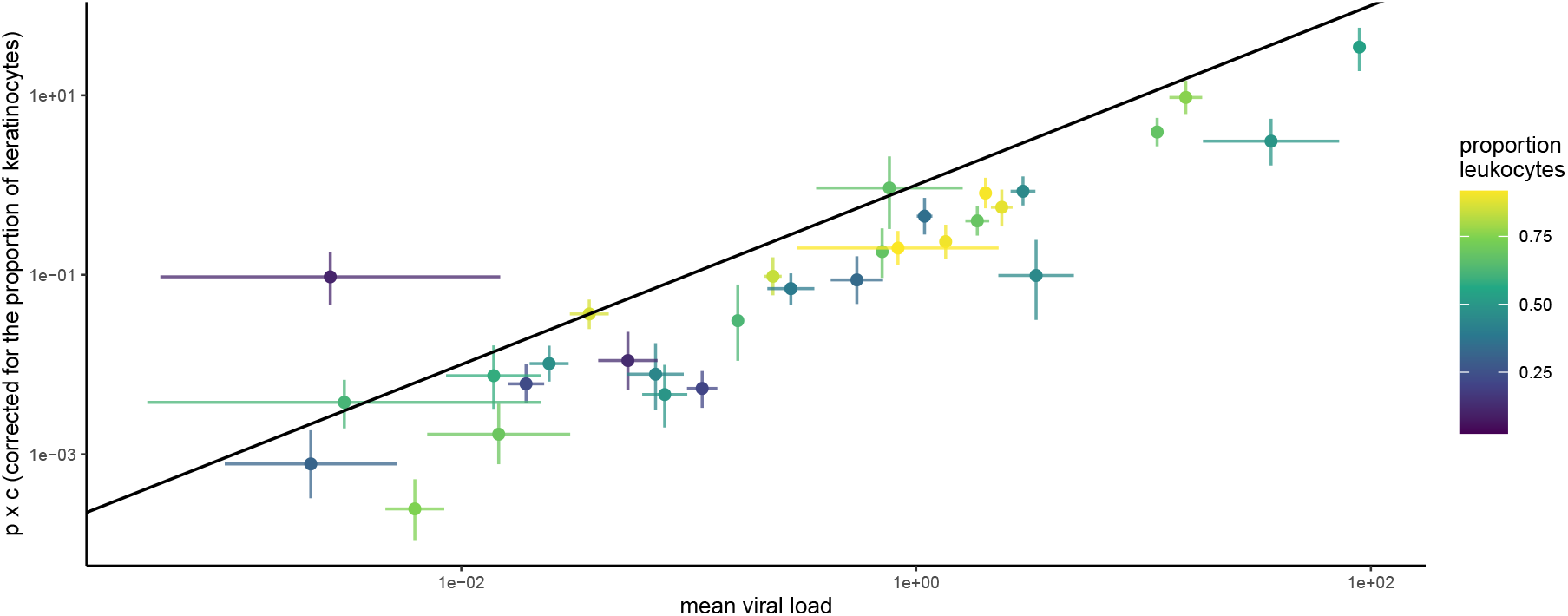
Mean viral load expected based on the limiting dilution assay as a function of the mean viral load measured by DNA column extraction. The expected virus load is the product of the estimates of *p, c*, and the proportion of keratinocytes. The colors indicate the proportion of leukocytes measured in the samples. The lines indicate the 95% CI of the estimates for the bulk sample (horizontal) and the 95% CrI for the limiting dilution estimates (vertical). The black line shows the expected 1:1 relationship.

